# Opioid overdose in public libraries: Results from a five state survey

**DOI:** 10.1101/2020.06.05.20123422

**Authors:** Rachel Feuerstein-Simon, Margaret Lowenstein, Roxanne Dupuis, Xochitl Luna Marti, Abby Dolan, Carolyn C. Cannuscio

## Abstract

**Introduction:** Public libraries are increasingly impacted by the overdose crisis. A 2017 survey of public librarians in the state of Pennsylvania revealed that 12% had reported an on-site overdose in the previous year. There are increasing public and private efforts to equip public libraries with the opioid overdose antidote, naloxone.

**Methods:** We conducted a cross-sectional web-based survey of all public library branches in Colorado, Connecticut, Florida, Michigan, and Virginia. Survey questions. We used descriptive statistics to report frequencies of responses and crude odds ratios were calculated to predict the dichotomized variable of endorsement of naloxone uptake.

**Results:** Library staff reported witnessing on-site alcohol (45%) and injection drug (14%) use in the past month. The one-year cumulative incidence (12% overall) of on-site overdose ranged from a low of 10% in MI, to a high of 17% in FL. Among libraries with on-site overdoses, a minority (21%) stocked naloxone, and 12% had administered naloxone. Overall, 11% of libraries stocked naloxone on-site. Although 24% of respondents reported attending at least one training regarding SUD in the past year, 91% wanted more training on the topic.

**Conclusions:** Public library staff routinely address issues related to substance use and overdose in their institutions. This work highlights the importance of including public libraries as part of a comprehensive public health strategy to address substance use-related morbidity and mortality in the U.S.

## Introduction

The United States is in the midst of a drug overdose crisis. As of 2017, over 7.5 million Americans over the age of 12 years have an illicit drug use disorder and over 14 million have alcohol use disorder.^1^ In addition, almost 70,000 Americans died from drug overdoses, with a majority of these deaths are due to opioids.^1^ Despite a modest reduction in overdose deaths in 2018, there continues to be an increase in deaths involving potent synthetic opioids like fentanyl.^2^ Further, there have been concerning increases in the rates of stimulant and alcohol use nationally.

Substance use and overdose in the U.S. is an ever-present reality in public spaces such as restaurants, parks, and public libraries. Public libraries are of particular interest as they are free and open to all, value privacy and equity, and frequently serve vulnerable patrons including those with mental health and substance use disorders.^3,4^ As such, library staff are increasingly feeling the effects of the overdose crisis, with 12% of Pennsylvania libraries reporting an on-site overdose in 2016.^5^

Public libraries across the country are beginning to develop policies and programs in response to substance use, such as installing containers for safe needle disposal.^6^ Others have actively taken on roles as first responders by stocking the opioid overdose antidote, naloxone, on-site and training their staff how to administer the medication.^7,8^ In 2018, the manufacturer of Narcan began offering two free doses of the medication to any interested public library branch in the U.S.^9^

Despite increasing reports of on-site drug use and overdose at libraries and a national effort to equip libraries with naloxone, questions remain regarding the burden of substance use in U.S. public libraries. In this context, we aimed to describe the scope of substance use and overdose in library settings using a cross-sectional survey of public libraries in five states. Results from the study will inform the public health community regarding the scope of the problem in public libraries, their capacity to respond, and assess future needs for resources and programming.

## Methods

### Study Design

Our team conducted a cross-sectional online survey of public libraries in Colorado, Connecticut, Florida, Michigan, and Virginia. These five states were chosen at random as the first phase of a planned study regarding the ways public libraries address community health needs.

### Survey Design and Administration

Our team, the Healthy Library Initiative (healthylibrary.org), developed a 25-item web-based cross-sectional survey. Items specifically focused on: 1) the frequency and outcomes of substance use and related emergencies in public libraries; 2) public librarian staff attitudes and concerns regarding substance use in their libraries and communities; 3) public library staff comfort and willingness to engage patrons who use drugs; and 4) implementation of library policies regarding substance use. Brief demographic information was also collected. These questions were administered as part of a larger National Public Library Community Health Survey deployed by our team. Survey development was informed by a qualitative study of public librarians conducted in 2015 and a single-state survey of public libraries conducted in 2017.^5,10^ The results presented in this paper are responses to substance use-related questions only.

### Survey administration

Survey invitations were emailed to 1,695 public library staff across Colorado, Connecticut, Florida, Michigan, and Virginia. We enumerated all public library branches in the five states, triangulating from multiple data sources including: 1) lists from state library directors, 2) membership rosters of the American Library Association, and 3) web searches in order to fully account for every branch in every state studied. We cross-referenced the databases we built with the Institute of Museum and Library Services (IMLS) Public Library Survey (PLS), a “census” of all public library branches in the U.S. The PLS does not include electronic contact information for library branches, only telephone numbers. Our team collected and verified that electronic contact information through a combination of web searches and telephone outreach in order to establish a known denominator of all libraries in the included states, with no “ghost libraries” (inactive contact addresses) in the pilot sample.

The survey was self-administered online by respondents using REDCap Version R v8.11.3, a secure online electronic data capture tool. Unique survey links were emailed to each respondent between August 2018 – May 2019. We allowed only one response per library. This was achieved by sending a unique survey link to one representative from each library. The survey asked participants to verify that they had direct patron contact. If the initial respondent did not have direct contact with patrons, they were asked to provide the email address of another contact at their library who met this criteria.

### Data Analysis

The survey data were analyzed using R v.1.1.45. We used descriptive statistics to report frequencies of responses and crude odds ratios were calculated to predict the dichotomized variable of endorsement of naloxone uptake. We calculated the relative odds of a library having acquired naloxone by the time of the survey, a dichotomous outcome. Our predictors were also dichotomous and were assessed at the same time as the outcome. Predictors included: strong vs. weak endorsement of the manufacturer campaign to donate naloxone to public libraries (top quartile vs. bottom quartile of score distribution, on a scale from 0-100); report of an overdose on-site in that library in the past year (yes/no); metro- vs. non-metro library location; county-level overdose mortality rate above or below the national median. Crude odds ratios were reported and future multivariate analyses are planned for the final manuscript.

We excluded respondents from analyses if they reported no direct patron contact or did not complete the survey. This study was approved by the Institutional Review Board at the University of Pennsylvania.

## Results

We received 356 total responses (response rate 21%).

Across the five states, 14% of responding library staff reported witnessing on-site injection drug use and 45% reported witnessing on-site alcohol use in the previous month (Table 1). There was a wide range by state in the proportion of respondents who reported witnessing on-site injection drug use from 36% in Colorado to a low of 7% in Connecticut.

**Table 1.** Reported frequency of on-site substance use in the previous month.

| Substance Type | CO<br>(n=36) | CT<br>(n=28) | FL<br>(n=65) | MI<br>(n=161) | VA<br>(n=66) | Total |
| --- | --- | --- | --- | --- | --- | --- |
| Injection drugs | 36% | 7% | 14% | 11% | 12% | 14% |
| Alcohol | 47% | 54% | 52% | 39% | 45% | 45% |

Across all states, 12% of respondents reported at least one on-site overdose in the previous year. The highest rate was in Florida (17%), followed by Colorado (14%), Virginia (12%), Connecticut (11%), and Michigan (10%).

Library staff reported a variety of policies and practices to address substance use and overdose in their libraries (Table 2). Overall, 11% of libraries stocked naloxone on-site. However, there was wide variation across states, ranging from 33% in Colorado, to 0% in Florida. Rates of naloxone uptake among libraries with on-site overdoses (21%) were higher than the overall average. Further, 12% of libraries that had experienced an on-site overdose had administered naloxone in the previous year. We saw lower rates of implementation of other practices, including formal guidelines for intervening when people used substances in the library (10%), restroom monitoring or time limits (7%), and safe needle disposal (6%) with some variability across states.

**Table 2.** Library policies and practices to address on-site substance use.

| Policy | CO | CT | FL | MI | VA | Total |
| --- | --- | --- | --- | --- | --- | --- |
| Naloxone (Narcan) stocked on-site in the library | 33% | 14% | 0% | 8% | 17% | 11% |
| Restroom monitoring or time limits | 6% | 7% | 3% | 10% | 5% | 7% |
| Safe needle disposal facilities | 33% | 4% | 3% | 2% | 2% | 6% |
| Formal guidelines for intervening when people are using substances in the library | 14% | 4% | 12% | 7% | 14% | 10% |

All four dichotomous predictors in Table 3 were associated with increased naloxone uptake by the library. The strongest association was observed for library location in a metro vs. non-metro area, followed by prior overdose in the library. Higher county-level overdose death rate was also associated with increased naloxone uptake, as was the individual librarian’s endorsement of a manufacturer-sponsored naloxone distribution effort--which is conceivably a modifiable lever for intervention to increase institutional naloxone uptake. Tests of significance will be performed in the final manuscript. These numbers were used to guide power estimates for planned studies.

**Table 3.** Predictors of naloxone update in public libraries.

| Exposure | Odds Ratio |
| --- | --- |
| Librarian endorses concept of naloxone in libraries (high vs. lower) | 1.7 |
| Prior overdose in library (yes/no) | 3.1 |
| Library located in metro vs. non-metro area | 4.6 |
| County-level overdose mortality (above vs. below the median) | 2.4 |

We also queried library staff regarding the frequency patrons’ request assistance with finding drug and alcohol treatment services. An average of 22% of library staff reported assisting patrons with finding drug and alcohol treatment services, ranging from a high of 36% in Florida to a low of 14% in Connecticut. Finally, Although 24% of respondents reported attending at least one training regarding SUD in the past year, 91% wanted more training on the topic.

## Discussion

In this five-state survey of public libraries, we found that alcohol and drug use in libraries was commonly reported. 12% of public libraries reported an on-site drug overdose, yet there was relatively low uptake of naloxone and other practices to mitigate the impacts of drug and alcohol use.^5^ We did find interest among librarians in receiving additional training to address these topics, suggesting that library staff may be recognizing the potential role of libraries in meeting the needs of those struggling with addiction.

In particular, we found that naloxone uptake remained low in states with a high burden of overdose morbidity and mortality. For example, Florida ranks 16th nationwide among states for overdose death rates, yet none of the 65 participating libraries from Florida stocked naloxone at the time of this study.^1^ Conversely, uptake was high in Colorado where overdose death rates are lower. Nonetheless, data from this study indicate that high levels of substance use, including injection drug use, in libraries, aligning with national data showing that no state or city is exempt from the effects of addiction. Since drug use is widespread and we cannot currently predict with precision in which libraries overdoses will occur, it is vital to increase the adoption of naloxone and overdose reversal trainings in public libraries across the county.

In addition to implementing naloxone and overdose reversal trainings in libraries, library staff would likely benefit from further training and thoughtfully formulated policies to support people who use drugs (PWUD). For example, in every participating state, less than 10% of libraries that they have routine monitoring of restrooms. While this type of policy could be perceived as unwanted surveillance, it also could provide a degree of supervision and protection for PWUD, in order to recognize and intervene rapidly in the event of an overdose. Furthermore, library staff are forced to improvise when interacting with all library visitors, including PWUD. They could benefit from training in de-escalation of conflict and in the principles of harm reduction. Central to any training, should be efforts to destigmatize drug use.

Further, library policies and staff training should recognize the broader challenges of substance use and addiction, including high rates of alcohol use. Trainings for library staff and resources for the public should address the underlying drivers of alcohol and substance use and addiction, including trauma. Libraries across the country are beginning to experiment with trauma-informed service delivery as well as on-site provision of social work services or connection to community-based services.

This study has several limitations. Importantly, we have surveyed libraries in only five states. These states were selected at random with the intention of building the sample over time to include libraries throughout the U.S. In addition, the libraries that participated may reflect those differentially engaged in addressing the substance use concerns of their patrons and communities. The response rate to this electronic survey (20%) was lower than the 80% response rate we have obtained from a statewide telephone survey of librarians (manuscript in preparation), indicating that telephone outreach may well be a superior tool for reaching this important group.

## Conclusion

Public library staff routinely address issues related to substance use and overdose in their institutions. These findings indicate a need for continued inquiry into the ways libraries nationally are impacted by the overdose crisis. This work also highlights the importance of including public libraries as part of a comprehensive public health strategy to address substance use-related morbidity and mortality.

## Data Availability

Due to privacy concerns of public librarians, the dataset cannot be made available.

